# In-hospital cardiac arrest in Intensive Care Unit versus non-Intensive Care Unit patients with COVID-19. A systematic review and meta-analysis

**DOI:** 10.1101/2021.05.08.21256885

**Authors:** Georgios Mavrovounis, Maria Mermiri, Athanasios Chalkias, Vishad Sheth, Vasiliki Tsolaki, Konstantinos Gourgoulianis, Ioannis Pantazoulos

## Abstract

**Aim:** To estimate the incidence of in-hospital cardiac arrest (IHCA) and return of spontaneous circulation (ROSC) in COVID-19 patients, as well as to compare the incidence and outcomes of IHCA in Intensive Care Unit (ICU) versus non-ICU patients with COVID-19.

**Methods:** We systematically reviewed the PubMed, Scopus and clinicaltrials.gov databases to identify relevant studies.

**Results:** Eleven studies were included in our study. The pooled prevalence/incidence, pooled odds ratios (OR) and 95% Confidence Intervals (95% CI) were calculated, as appropriate. The quality of the included studies was assessed using appropriate tools. The pooled incidence of IHCA in COVID-19 patients was 7% [95% CI: 4 – 11%; P < 0.0001] and 44% [95% CI: 30 – 58%; P < 0.0001] achieved ROSC. Of those that survived, 58% [95% CI: 42 – 74%; P < 0.0001] had a good neurological outcome (Cerebral Performance Category 1 or 2) and the mortality at the last follow-up was 59% [95% CI: 37 – 81%; P < 0.0001]. A statistically significant higher percentage of ROSC [OR (95% CI): 5.088 (2.852, 9.079); P < 0.0001] was found among ICU patients versus those in the general wards.

**Conclusion:** The incidence of IHCA amongst hospitalized COVID-19 patients is 7%, with 44% of them achieving ROSC. Patients in the ICU were more likely to achieve ROSC than those in the general wards, however the mortality did not differ.

**What this paper adds:** Section 1: What is already known on this subject

- Mortality in COVID-19 patients ranges between 20% and 40%.
- it has been reported that patients with COVID-19 have a high incidence of IHCA and higher mortality.
- This paper aimed to calculate the proportion of COVID-19 patients who experience IHCA and their outcome, as well as compare the outcome of IHCA between ICU and non-ICU patients.

Section 2: What this study adds

- Approximately 7% of hospitalized COVID-19 patients suffer from IHCA and 44% of those achieve ROSC.
- The rate of ROSC was higher in ICU patients, but the rate of mortality did not differe between ICU and non-ICU patients.

## Introduction

The Coronavirus disease 2019 (COVID-19) pandemic has substantially impacted healthcare worldwide, causing more than 2 million deaths since the original outbreak in December 2019 [1]. The clinical syndrome caused by the SARS-CoV-2 virus ranges from mild respiratory infection to pneumonia, acute respiratory distress syndrome (ARDS), multiorgan failure and death [2]. There have been reports that mortality ranges between 20% in patients with mild symptoms and 40% in critically-ill patients [3,4]. Moreover, it has been reported that patients with COVID-19 have a higher incidence of In-Hospital-Cardiac-Arrest (IHCA) [5] and worse outcomes [6]. However, few studies have attempted to estimate the incidence and outcomes of IHCA in COVID-19 patients so far [7].

The incidence of IHCA in the pre-COVID-19 era varied significantly between studies [8,9]. Patients with witnessed IHCA and with ventricular fibrillation (VF) or pulseless ventricular tachycardia (pVT) have a higher incidence of return of spontaneous circulation (ROSC) [10]. However, it has been showcased that most IHCAs present with non-shockable rhythms, namely asystole or pulseless electrical activity (PEA) [11], with approximately 20-25% of patients surviving to hospital discharge [11]. The majority of IHCAs occur in the Intensive Care Unit (ICU), where the existence of advanced monitoring equipment and highly trained personnel leads to higher ROSC rates [12].

The aim of the current systematic review and meta-analysis is to estimate the incidence or IHCA and ROSC in COVID-19 patients, as well as to compare the incidence and outcomes of IHCA in ICU versus non-ICU patients with COVID-19.

## Methods

The protocol for the current systematic review and meta-analysis was registered on the International prospective register of systematic reviews (PROSPERO ID number: CRD42020226262) and is available in full on: https://www.crd.york.ac.uk/prospero/display_record.php?RecordID=226262.

### Patient and Public Involvement Statement

No patient and public involvement statement is required for this study, as this is a systematic review of the literature.

### Literature search

Two of the authors (M.G. and M.M.) individually performed a database search of the PubMed (MEDLINE), Scopus and clinicaltrials.gov databases. The following keywords were used and combined with the Boolean operators “AND” and “OR” as appropriate: “severe acute respiratory syndrome coronavirus 2”, “ncov”, “2019 ncov”, “covid 19”, “sars cov 2”, “coronavirus”, “cov”, “heart arrest”, “IHCA”, “in-hospital cardiac arrest”, “in-hospital cardiopulmonary arrest”. The last literature search was performed on February 24^th^, 2021. The exact search algorithms for all three databases are available in Appendix A. We reviewed the reference lists of the retrieved articles to identify additional original studies that fulfilled our criteria.

### Inclusion and exclusion criteria

Our inclusion criteria were: (1) Randomized Controlled Trials (RCTs), prospective cohort studies, retrospective cohort studies, short communications, letters to the editor and conference abstracts with original data, (2) reporting on the outcomes of adult COVID-19 patients with IHCA, (3) treated in the ICU or general wards. Only studies (4) written in English were included.

We excluded studies (1) on pediatric patients, (2) patients with unconfirmed COVID-19 diagnosis and (3) studies on patients with out-of-hospital cardiac arrest. Case reports were also excluded. Finally, (4) studies in languages other than English were also excluded.

### Data extraction

Two independent investigators (M.G. and M.M.) used a standardized data form to execute the data extraction process. The following data were extracted: First author’s name, Year of publication, Mean/Median age of the patients, Male/Female ratio, Number of hospitalized COVID-19 patients (if available the number of patients in the ICU and in general wards was noted), Number of hospitalized COVID-19 patients that experienced IHCA (if available the number of patients in the ICU and in general wards was noted), Number of IHCA patients that achieved ROSC (if available the number of patients in the ICU and in general wards was noted), arrest rhythm, mortality at last follow-up (also, mortality at 30-day and 6 months of follow-up, if available), neurological outcome at the last available follow-up for each study, etiology of witnessed arrest, comorbidities and length of follow-up. Any discrepancies between the reviewers were resolved by two other investigators (P.I., A.C). For missing data we tried to contact the authors and asked them to provide the necessary raw data.

### Outcomes

The outcome of the study was two-fold. Firstly, we aimed to combine all the available studies in patients with IHCA and a COVID-19 diagnosis by implementing a random effects model meta-analysis using event rates and 95% Confidence Intervals (CI), to calculate the following: The percentages of hospitalized Covid-19 patients 1) that experienced an episode of IHCA, 2) achieved ROSC, (3) with IHCA that did not survive at the last follow-up (mortality), (4) with ROSC that died at the last follow-up, (5) with a good neurological outcome at the last available follow-up for each study [as defined by a Cerebral Performance Category (CPC) score of 1 or 2], (6) experiencing IHCA, previously diagnosed with various comorbidities (e.g. diabetes mellitus, arterial hypertension, coronary artery disease, malignancy, chronic kidney disease, chronic lung diseases).

Furthermore, when enough data were available in the literature, we aimed to compare patients that experienced IHCA while being treated in the ICU versus those in the general wards, regarding: (1) IHCA rate (= number of arrests/ number of COVID-19 patients), (2) ROSC rate (= number of patients that achieved ROSC/ number of arrests), (3) death rate at last follow-up (= number of deaths/ number of arrests), (4) death rate at last follow-up in patients that achieved ROSC, (5) death rate at 30-day follow-up (= number of deaths at 30 days/ number of arrests), (6) death rate at 6 months (= number of deaths at 6 months/ number of arrests), (7) rates of patients with a good neurological outcome (number of patients with CPC 1 or 2/ number of patients that achieved ROSC).

### Risk of bias assessment

As all the included studies were observational, the Methodological index for non-randomized studies (MINORS) [13] assessment tool was used to assess their quality. MINORS is a 12-item tool designed to assess the methodological quality of comparative and non-comparative non-randomized studies.

### Statistical analysis

All statistical analyses were performed in Review Manager (RevMan) Version 5.3 software (The Nordic Cochrane Centre, The Cochrane Collaboration, Copenhagen, Denmark (http://tech.cochrane.org/revman)) and OpenMeta[Analyst] [14].

Proportional (event rate) meta-analyses were performed by calculating the pooled event rates and 95% CI. The comparisons between ICU versus non-ICU/general ward patients were performed by calculating the pooled odds ratio (OR) and 95% CI. The significance was set at P<0.05. The statistical significance of the OR was determined by the use of the Z test.

Cochran’s Q and I^2^ indices were calculated to estimate the statistical heterogeneity of the studies. A random effects model was applied when I^2^>50% and/or P_Q_<0.10, otherwise a fixed effects model was implemented [15]. Funnel plots and the Egger’s test were used to assess for publication bias; P values less than 0.05 indicate significant publication bias [16–18].

The PRISMA guidelines for reporting systematic reviews and meta-analyses were applied (Appendix B).

## Results

### Selection of the included studies

Our electronic database search resulted in 373 articles, after removal of the duplicates. Following the title and abstract review, we identified 27 potentially relevant articles. The full texts of those articles were thoroughly reviewed, resulting in 11 studies that fulfilled our inclusion criteria and were included in our systematic review. The flowchart for study selection according to the PRISMA guidelines is presented in Fig. 1.

**Figure 1:**
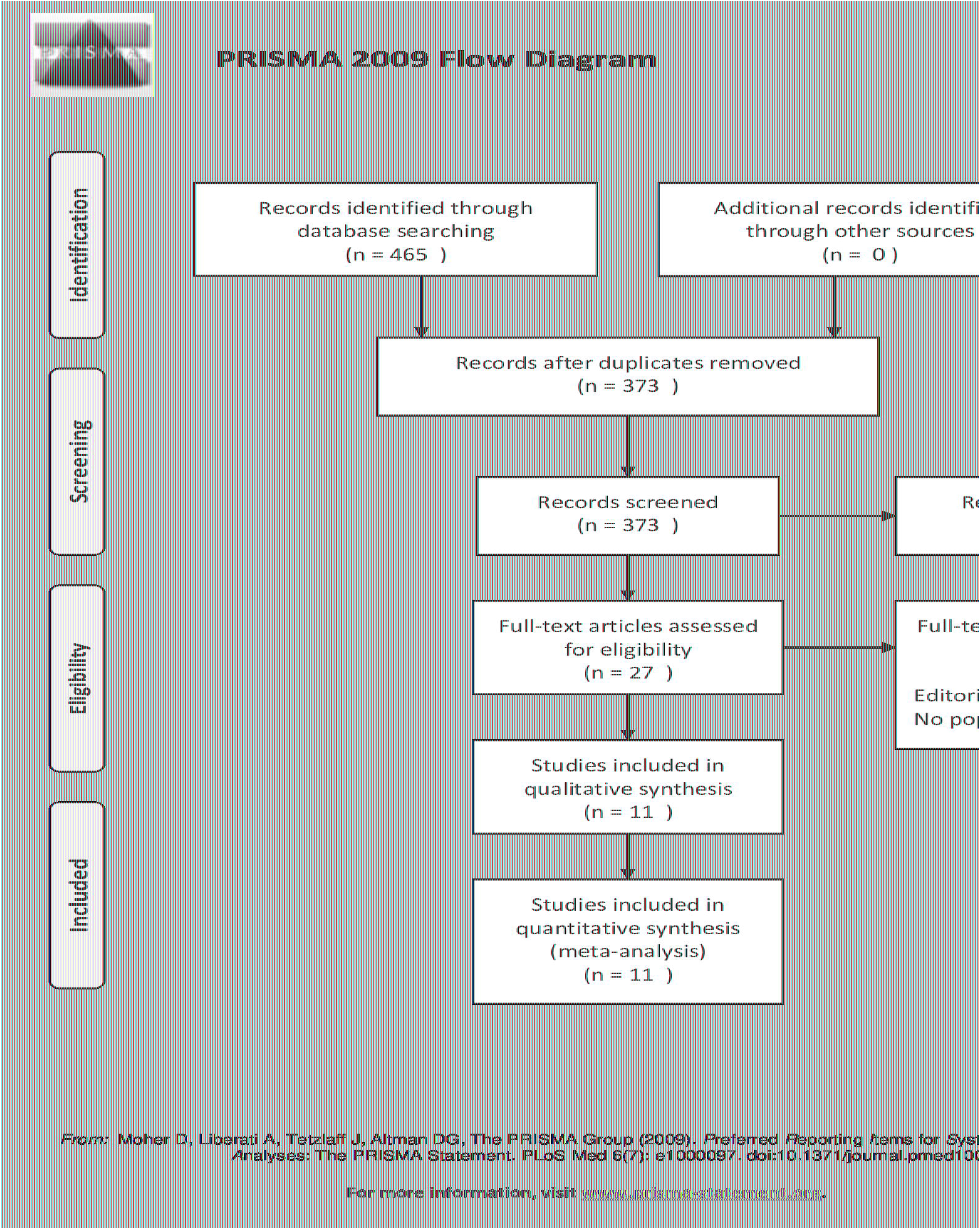
PRISMA flowchart for study selection

### Study and patient characteristics

All studies included were observational and were published in 2020-2021. Nine studies provided data regarding IHCA patients treated in the general wards, while 2 studies included patients in the ICU. Table 1 summarizes the study characteristics, including age, male/female ratio, arrest rhythm and etiology, patients’ comorbidities etc.

**Table 1:**
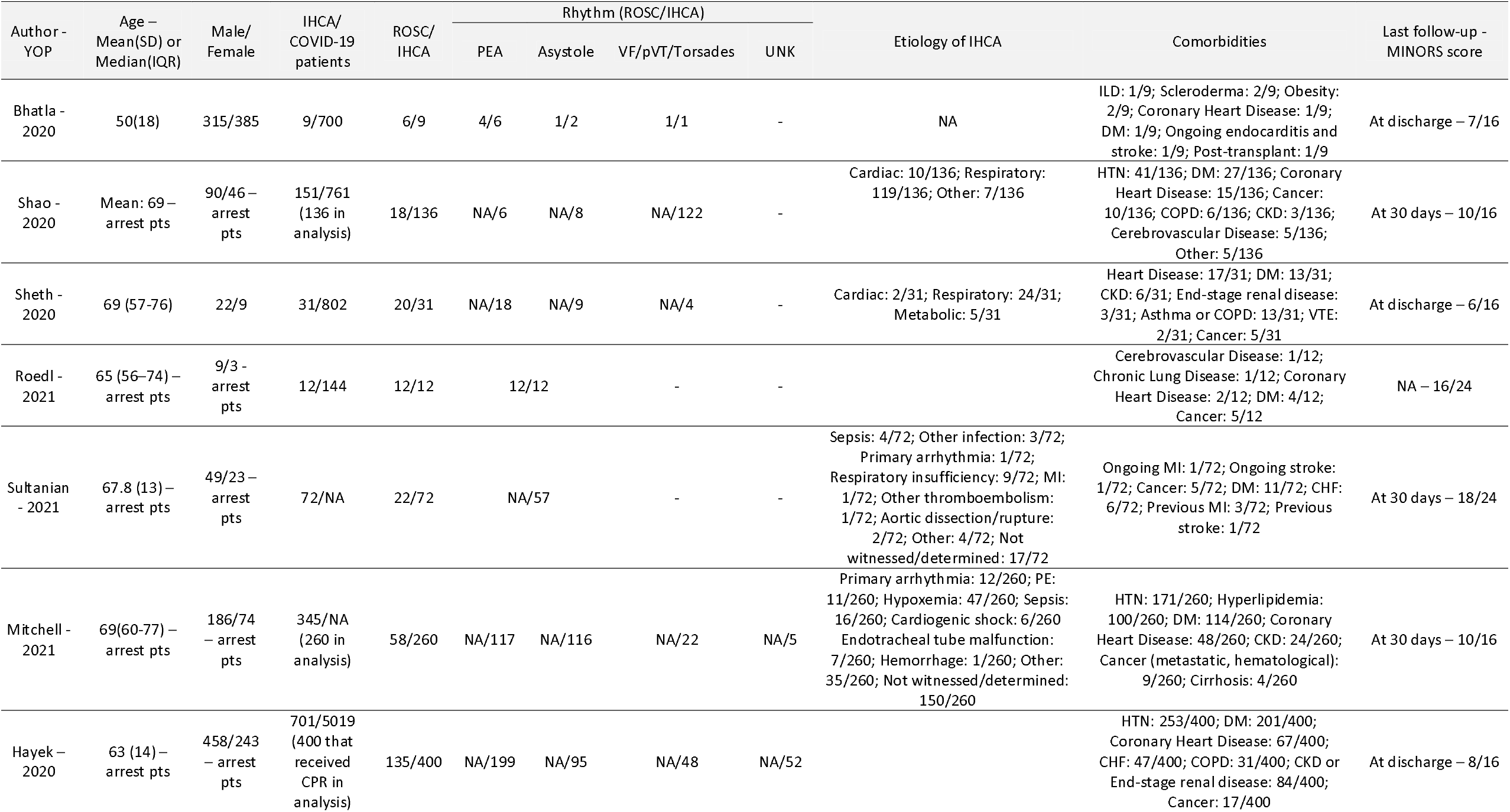
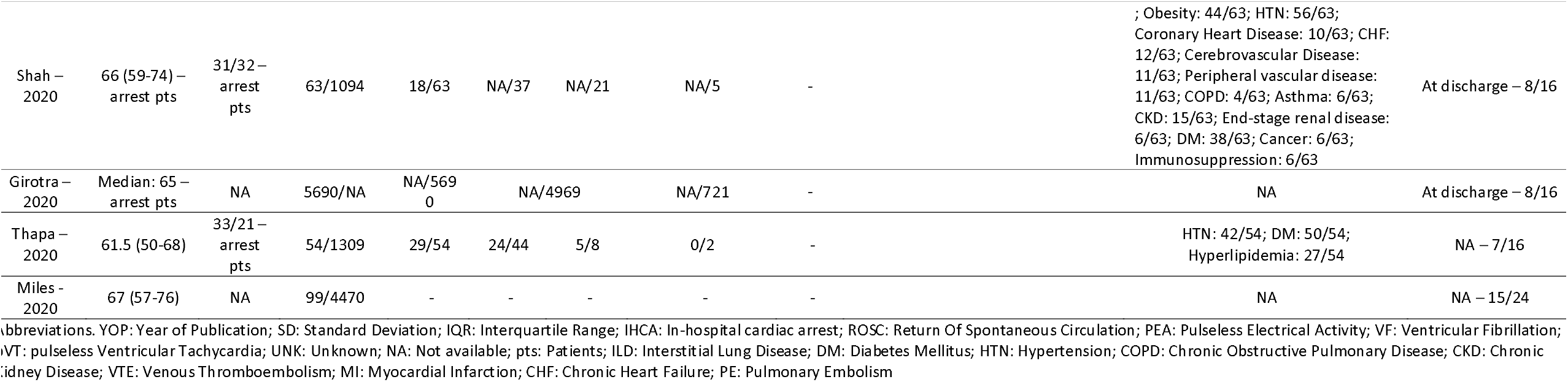
Table presenting the main characteristics of the included studies, along with the results of the MINORS tool.

### Publication bias and risk of bias assessment

The visual inspection of the funnel plots for each analysis, as well as the Egger’s test didn’t reveal significant asymmetry for most of the analyses (significant only for cancer prevalence). All funnel plots are presented in Appendix C; the results of the Egger’s test are available in Appendix D.

The results of the risk of bias assessment according to the MINORS tool are presented in Table 1.

### In-hospital cardiac arrests and ROSC in COVID-19 patients

Eight out of the 11 included studies provided data on the number of IHCA events amongst hospitalized COVID-19 patients. The pooled incidence of IHCA in COVID-19 patients was 7% [95% CI: 4 – 11%; P < 0.0001] (Fig. 2A).

**Figure 2:**
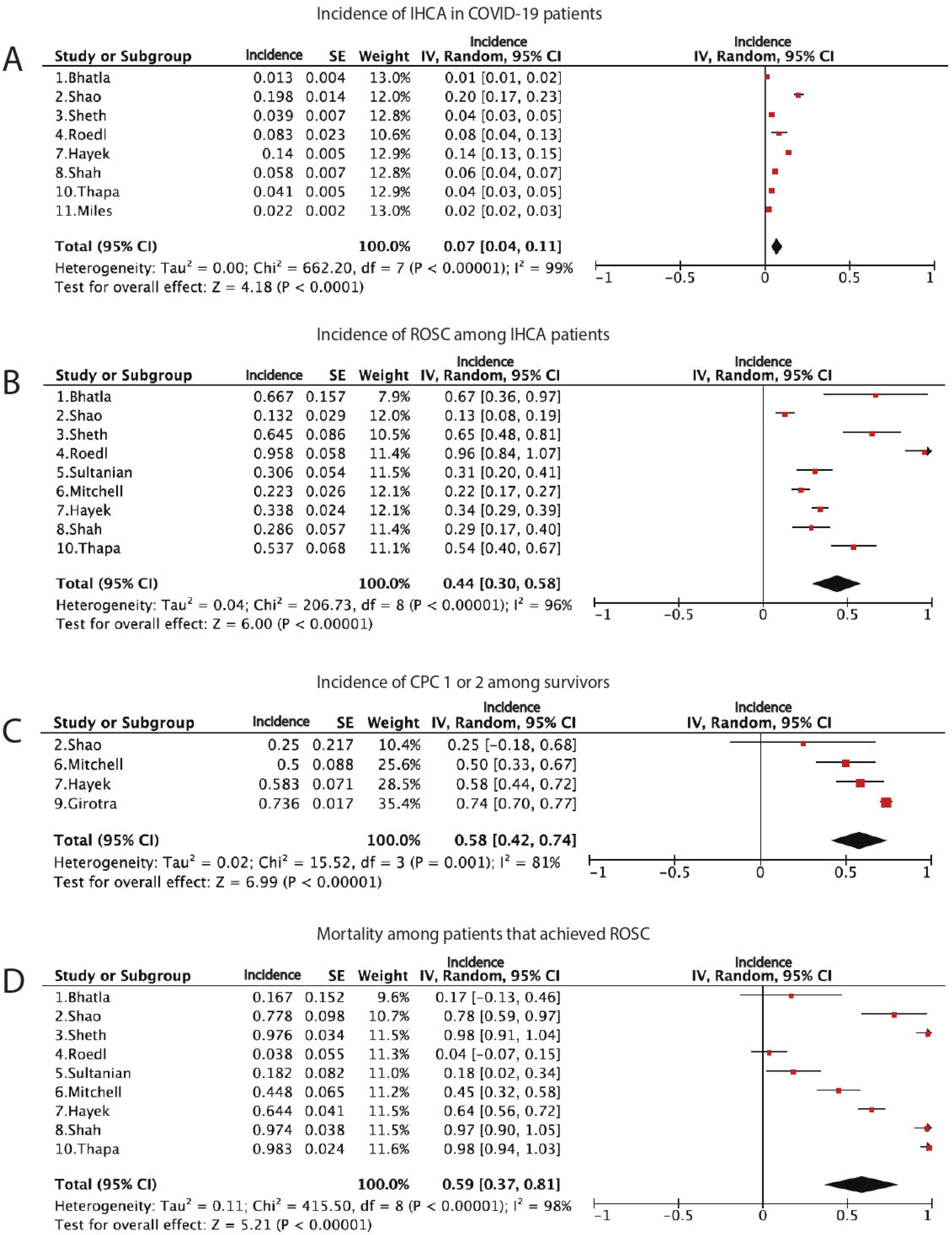
Forest plots presenting the results of proportional meta-analyses regarding the (A) number of In-Hospital cardiac arrests (IHCA) amongst patients with COVID-19, (B) number of patients that achieved ROSC after an episode of IHCA, (C) number of patients with a Cerebral Performance Category score of 1 or 2 amongst survivors, (D) number of patients that died at the last follow-up amongst those that achieved ROSC.

Nine out of the 11 included studies provided data on the number of COVID-19 patients that achieved ROSC after an episode of IHCA. The pooled incidence of ROSC in COVID-19 patients with IHCA was 44% [95% CI: 30 – 58%; P < 0.00001] (Fig. 2B).

### Cerebral Performance Category and mortality

Four out of the 11 included studies provided data on the number of patients with a good neurological outcome (CPC 1 or 2) at the last follow-up. The pooled incidence of CPC 1 or 2 among the survivors was 58% [95% CI: 42 – 74%; P < 0.00001] (Fig. 2C).

Nine out of the 11 included studies provided data on mortality at the last follow-up. The pooled mortality at the last available follow-up for each study amongst those that achieved ROSC was 59% [95% CI: 37 – 81%; P < 0.00001] (Fig. 2D).

### Arrest rhythms in COVID-19 patients

The pooled incidence of PEA in COVID-19 patients with IHCA was 50% [95% CI: 28 – 71%; P < 0.0001] (Fig. 3A). The pooled incidence of asystole in COVID-19 patients with IHCA was 23% [95% CI: 13 – 33%; P < 0.0001] (Fig. 3B). The pooled incidence of both non-shockable rhythms in COVID-19 patients with IHCA was 77% [95% CI: 63 – 91%; P < 0.0001] (Fig. 3C). The pooled incidence of pVT/V-fib/Torsades (shockable rhythms) in COVID-19 patients with IHCA was 18% [95% CI: 8 – 29%; P = 0.0004] (Fig. 3D).

**Figure 3:**
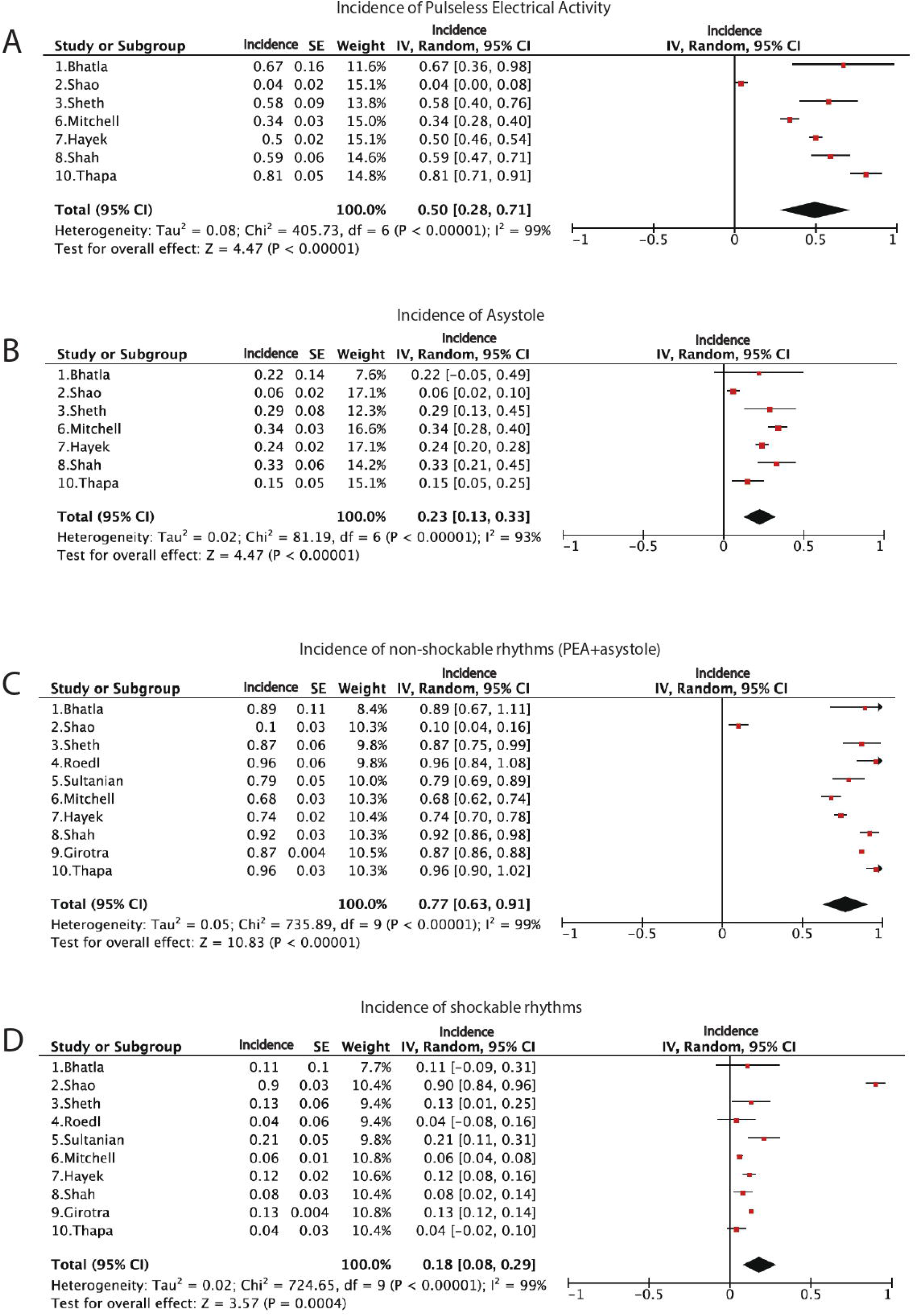
Forest plots presenting the results of proportional meta-analyses regarding the presenting arrest rhythms. (A) Pulseless Electrical Activity (PEA), (B) Asystole, (C) Non-shockable rhythms (PEA + Asystole), (D) Shockable rhythms (Pulseless Ventricular Tachycardia, Ventricular Fibrillation, Torsades de pointes).

### Patients’ Comorbidities

Data on coronary artery disease were available in 7 out of the 11 included studies. The pooled prevalence of coronary artery disease amongst all COVID-19 patients that experienced an episode of IHCA was 13% [95% CI: 8 – 18%; P < 0.00001].

Data on arterial hypertension were available in 5 out of the 11 included studies. The pooled prevalence of arterial hypertension amongst all COVID-19 patients that experienced an episode of IHCA was 65% [95% CI: 48 – 82%; P < 0.00001].

Data on diabetes mellitus were available in 9 out of the 11 included studies. The pooled prevalence of diabetes mellitus amongst all COVID-19 patients that experienced an episode of IHCA was 41% [95% CI: 24 – 59%; P < 0.00001].

Data on cancer were available in 7 out of the 11 included studies. The pooled prevalence of cancer amongst all COVID-19 patients that experienced an episode of IHCA was 6.3% [95% CI: 3.5 – 9.1%; P < 0.001].

Data on Chronic Lung Diseases were available in 6 out of the 11 included studies. The pooled prevalence of Chronic Lung Diseases amongst all COVID-19 patients that experienced an episode of IHCA was 9.1% [95% CI: 3.9 – 14.3%; P < 0.001].

Data on Chronic Kidney Diseases were available in 5 out of the 11 included studies. The pooled prevalence of Chronic Kidney Diseases amongst all COVID-19 patients that experienced an episode of IHCA was 14.3% [95% CI: 5.4 – 23.2%; P = 0.002].

### ICU vs. Non-ICU patients

Studies that compared the outcomes of patients treated in the ICU versus those hospitalized in the general wards were limited. Five studies compared the ROSC amongst IHCA patients in the ICU versus those in the general wards and 3 studies compared the number of deaths amongst IHCA patients in the ICU versus those in the general wards.

Patients in the ICU were statistically more likely to achieve ROSC than those in the general wards [OR (95% CI): 5.088 (2.852, 9.079); P < 0.0001] (Fig. 4A). No statistical difference was found in mortality amongst those that suffered an episode of IHCA in the ICU versus those in the general wards [OR (95% CI): 0.221 (0.037, 1.338); P = 0.1] (Fig. 4B).

**Figure 4:**
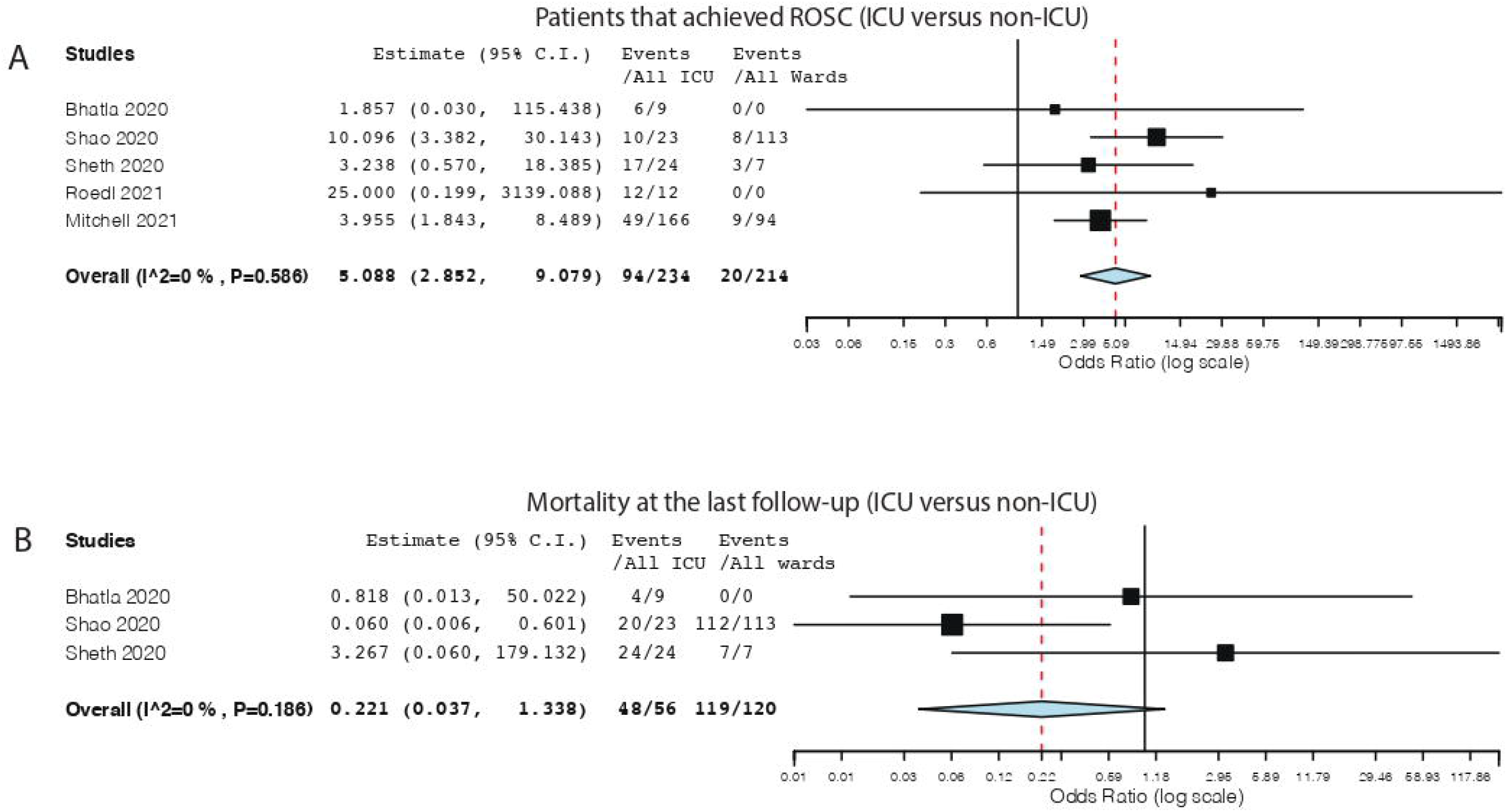
Forest plots presenting the results of comparison between ICU and non-ICU patients that (A) achieved ROSC after In-hospital cardiac arrest (IHCA), (B) died at the last follow-up (amongst those that had an episode of IHCA).

## Discussion

Our systematic review and meta-analysis demonstrates that the incidence of IHCA amongst hospitalized COVID-19 patients is 7%, with 44% of them achieving ROSC. Patients in the ICU were statistically more likely to achieve ROSC than those in the general wards, however no statistical difference was found in mortality. To our knowledge, this is the first systematic review and meta-analysis that compared ROSC and mortality rates between ICU and non-ICU COVID-19 patients at the time of the arrest.

The incidence of IHCA during the COVID-19 pandemic has been reported to be higher than usual (1-5/1000 hospital admissions), at 6.6 per 1000 hospital admissions [5,11]. This can be explained by the fact that COVID-19 patients are at high risk due to rapidly worsening respiratory failure, eventually leading to IHCA if not promptly treated. Furthermore, direct effects of the virus on the cardiovascular system might contribute to the increased risk for IHCA [5,19–21]. In the literature, the incidence of IHCA amongst COVID-19 patients ranges between 1.3% and 19.8%. Our study identified a pooled incidence of 7%.

Since the beginning of the pandemic, various authors have suggested that cardiac arrest and COVID-19 is a lethal combination with poor outcomes [22,23], mainly due to delays in initiating resuscitative procedures. Indeed, early studies from China [24] reported poor resuscitation outcomes and an alarmingly high percentage of asystole in patients with IHCA and COVID-19 infection. According to Mitchell et al. [25], such a high percentage of asystole might indicate significant delay in recognizing IHCA events, resulting in low ROSC and survival rates. Studies published later in the course of the pandemic [25–27], reported lower percentages of asystole and higher ROSC rates. Our results regarding the IHCA rhythm and ROSC reflect those later reports and are in-line with the results of previous studies in non-COVID-19 IHCA patients [28,29]. Although most IHCAs occur in general wards [8], the IHCA location has shifted towards the ICU during the COVID-19 pandemic (5). The high ROSC rate of 44% found in our study might be explained by the higher percentages of IHCA occurring in the ICU, where the arrest is usually identified and treated more promptly [5].

During the course of the pandemic, it has been established that patients with comorbidities such as diabetes mellitus, arterial hypertension, chronic kidney disease etc., are at a higher risk of severe COVID-19 infection [30]. Most authors reporting on the comorbidities of COVID-19 patients with IHCA, agree that these patients often suffer from cardiovascular diseases and diabetes mellitus [24,26,27]. In our analysis, arterial hypertension was the leading comorbidity with a pooled prevalence of 65%. Other commonly reported comorbidities were diabetes mellitus, chronic kidney disease, coronary artery disease, chronic lung disease and malignancy.

In the literature, the reported mortality amongst COVID-19 patients with IHCA achieving ROSC varies widely. Some authors report mortality rates as high as 80% - 90% [6,24,27,31], while others report distinctly lower rates [5]. Our analysis showed a pooled mortality of 59% among those that achieved ROSC, at the last available follow-up for each study. The disunity on the reported mortality among studies can be explained by several factors. Firstly, the percentage of severely ill patients treated with mechanical ventilation and/or renal replacement therapy at the time of the arrest, differed between studies. Secondly, not all institutions implemented a strategy of early ICU admission to enable meticulous monitoring and early identification of deterioration.

It is well-established that the location of the IHCA correlates with the outcome. Patients with IHCA in a monitored setting such as the ICU, are more likely to achieve ROSC and survive to hospital discharge [12,32–34]. It is not clear if this is also true in COVID-19 patients, as studies comparing patients in the ICU versus those in the general wards are scarce. Even though our analysis revealed a significantly higher occurrence of ROSC in the ICU versus non-ICU patients, it is important to note that this was not the case when deaths at the last available follow-up for each study were compared. The high mortality in the ICU is likely multifactorial. Many ICU patients receive mechanical ventilation, renal replacement therapy, and/or vasopressor support, and all these factors are associated with a poor outcome following IHCA [11]. Furthermore, the delayed admission of critically ill COVID-19 patients to ICU due to the paucity of available beds, which is well associated with increased mortality, may have an impact on the survival of post cardiac arrest COVID-19 patients. In a retrospective study of more than 50,000 patients, Chalfin et al. observed that critically ill patients who had more than a six-hour delay in ICU transfer had an increased ICU mortality [35].

Another important result was the high percentage of good neurological outcome among survivors. Half of the survivors presented a favorable neurological outcome (CPC 1 or 2). This result may provide an answer to the recently raised dilemma “to do or not to do CPR in COVID-19 patients?” [36]. Nowadays, with the majority of healthcare professionals have been vaccinated against COVID-19 and the adequacy of personal protective equipment, one can’t ignore the really high percentage of good neurological outcome among survivors of cardiac arrest. Thus, it seems that the risk-benefit balance for CPR has turned in favor of the benefit, and clinicians should keep a low threshold for performing CPR in COVID-19 patients experiencing cardiac arrest.

Our study has some limitations that should be acknowledged. Due to the COVID-19 pandemic being a recent event, there are limited data available in the literature, especially pertaining to the comparison between ICU versus non-ICU patients with IHCA. Furthermore, it is important to note that the included studies reported outcomes at different follow-up points. Some studies only included patients with IHCA treated in the ICU and didn’t analyze data concerning patients from the general wards. Finally, only observational studies were available in the literature.

## Conclusion

Our study demonstrates that the incidence of IHCA amongst hospitalized COVID-19 patients is 7% and that 44% of them achieve ROSC. Patients in the ICU were statistically more likely to achieve ROSC than those in the general wards, however no statistical difference was found in mortality.

## Data Availability

The data that support the findings of this study are available from the corresponding author (MM), upon reasonable request

## Abbreviations

COVID-19: (Coronavirus disease 2019)
ARDS: Acute Respiratory Distress Syndrome
IHCA: In-hospital Cardiac Arrest
VF: Ventricular Fibrillation
pVT: Pulseless Ventricular Tachycardia
ROSC: Return of Spontaneous Circulation
PEA: Pulseless Electrical Activity
ICU: Intensive Care Unit
CPC: Cerebral Performance Category

## Titles and Legends

**Appendix A:** Exact algorithms used for all databases.

**Appendix B:** PRISMA checklist for reporting systematic reviews and meta-analyses.

**Appendix C:** Funnel plots for investigation of publication bias. (A) In-hospital cardiac arrests (IHCA) amongst COVID-19 patients, (B) patients that achieved ROSC after an episode of IHCA, (C) patients with Pulseless Electrical Activity amongst those with IHCA, (D) patients with asystole amongst those with IHCA, (E) patients with non-shockable rhythms amongst those with IHCA, (F) patients with shockable rhythms amongst those with IHCA, (G) patients with a Cerebral Performance Category score of 1 or 2 amongst survivors, (H) number of deaths at the last follow-up amongst those that achieved ROSC, (I) patients with Coronary Artery Disease among those with IHCA, (J) patients with hypertension among those with IHCA, (K) patients with Diabetes Mellitus among those with IHCA, (L) patients with cancer among those with IHCA, (M) patients with Chronic Lung Diseases among those with IHCA, (N) patients with Chronic Kidney Diseases among those with IHCA, (O) patients that achieved ROSC in the ICU versus those in the general wards, (P) patients that died at the last follow-up amongst those that had an episode of IHCA in the ICU versus those in the general wards.

**Appendix D:** Results of the Egger’s test for all analyses.

