## Appendix A for "In-hospital cardiac arrest in Intensive Care Unit versus non-Intensive Care Unit patients with COVID-19. A systematic review and meta-analysis"

PubMed

("severe acute respiratory syndrome coronavirus 2"[Supplementary Concept] OR "severe acute respiratory syndrome coronavirus 2"[All Fields] OR "ncov"[All Fields] OR "2019 ncov"[All Fields] OR "covid 19"[All Fields] OR "sars cov 2"[All Fields] OR (("coronavirus"[All Fields] OR "cov"[All Fields]) AND 2019/11/01:3000/12/31[Date - Publication])) AND ("heart arrest"[MeSH Terms] OR "IHCA"[Title/Abstract] OR "in-hospital cardiac arrest"[Title/Abstract] OR "in-hospital cardiac arrest"[Title/Abstract] OR "in-hospital cardiopulmonary arrest"[Title/Abstract] OR "in-hospital cardiopulmonary arrest"[Title/Abstract])

### *Scopus*

### *( TITLE-ABS-KEY (* "severe acute respiratory syndrome coronavirus 2"  *OR*  "ncov"  *OR*  "2019 ncov"  *OR*  "covid 19"  *OR*  "sars cov 2"  *OR*  "coronavirus"  *OR*  "cov" *) )  AND  ( TITLE-ABS-KEY (* "heart arrest"  *OR*  "IHCA"  *OR*  "in-hospital cardiac arrest"  *OR*  "in-hospital cardiac arrest"  *OR*  "in-hospital cardiopulmonary arrest" *) )  AND  ( LIMIT-TO ( LANGUAGE ,*  "English" *) )*

Clinicaltrials.gov

Covid19 AND "cardiac arrest"
