## Supplementary figures and images for "In-hospital cardiac arrest in Intensive Care Unit versus non-Intensive Care Unit patients with COVID-19. A systematic review and meta-analysis"

### Appendix C

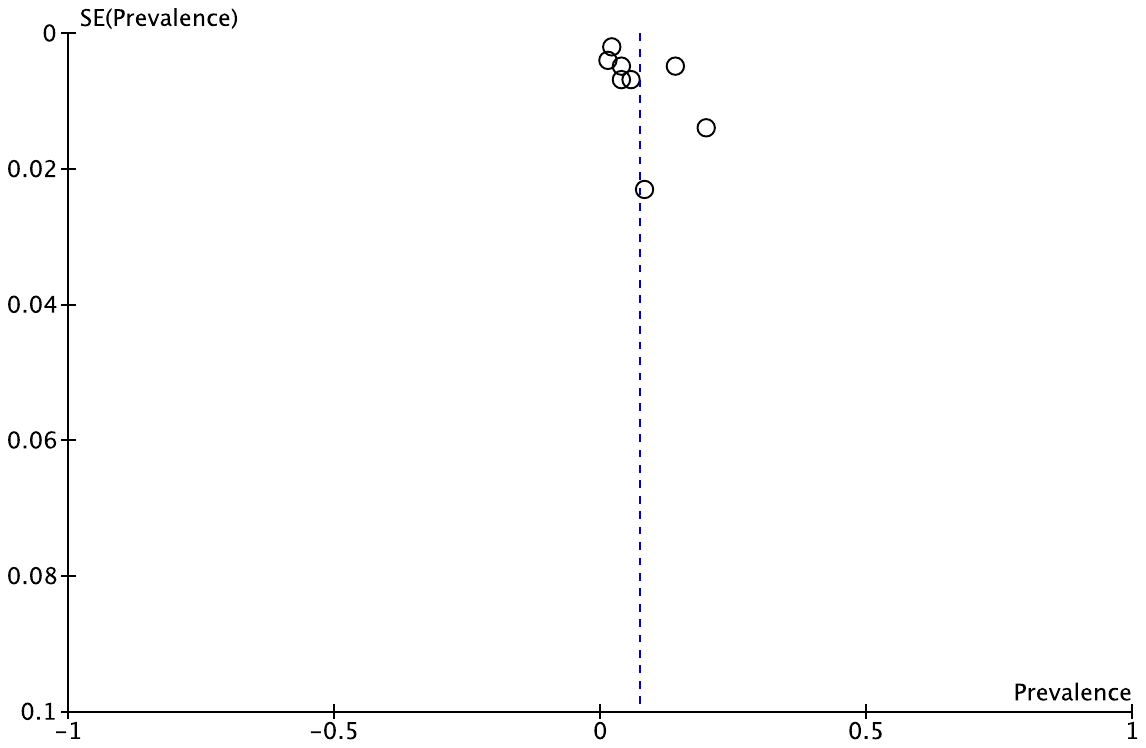
A.

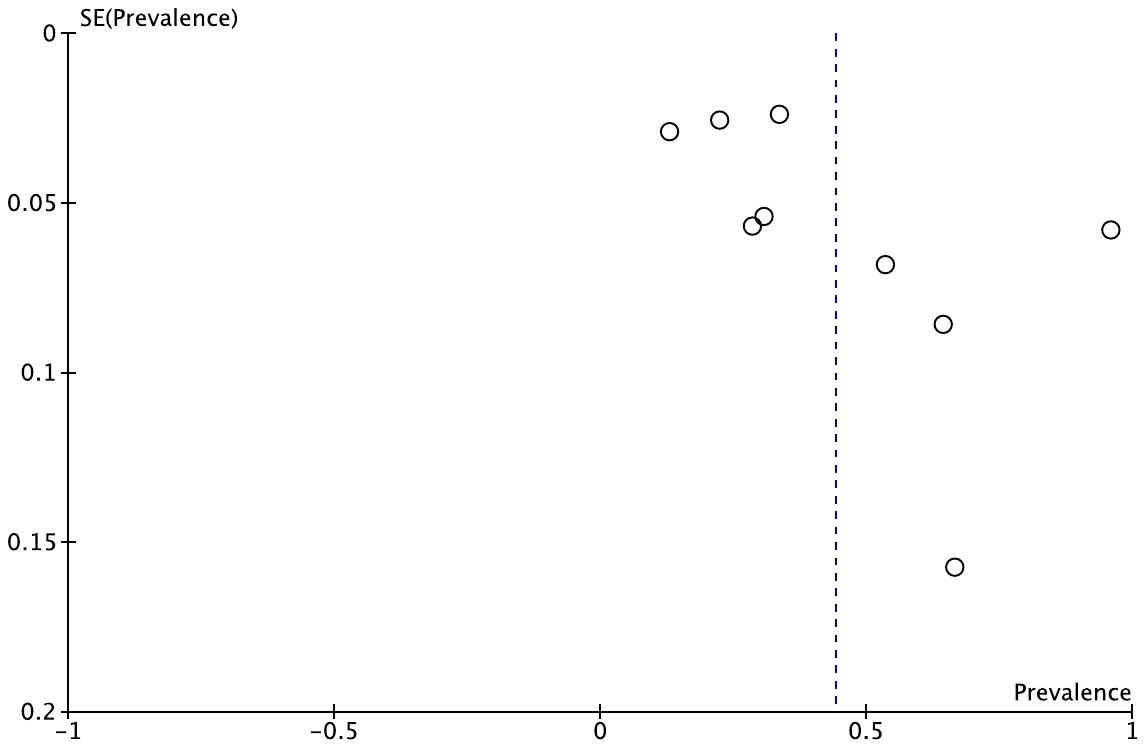
B.

C.

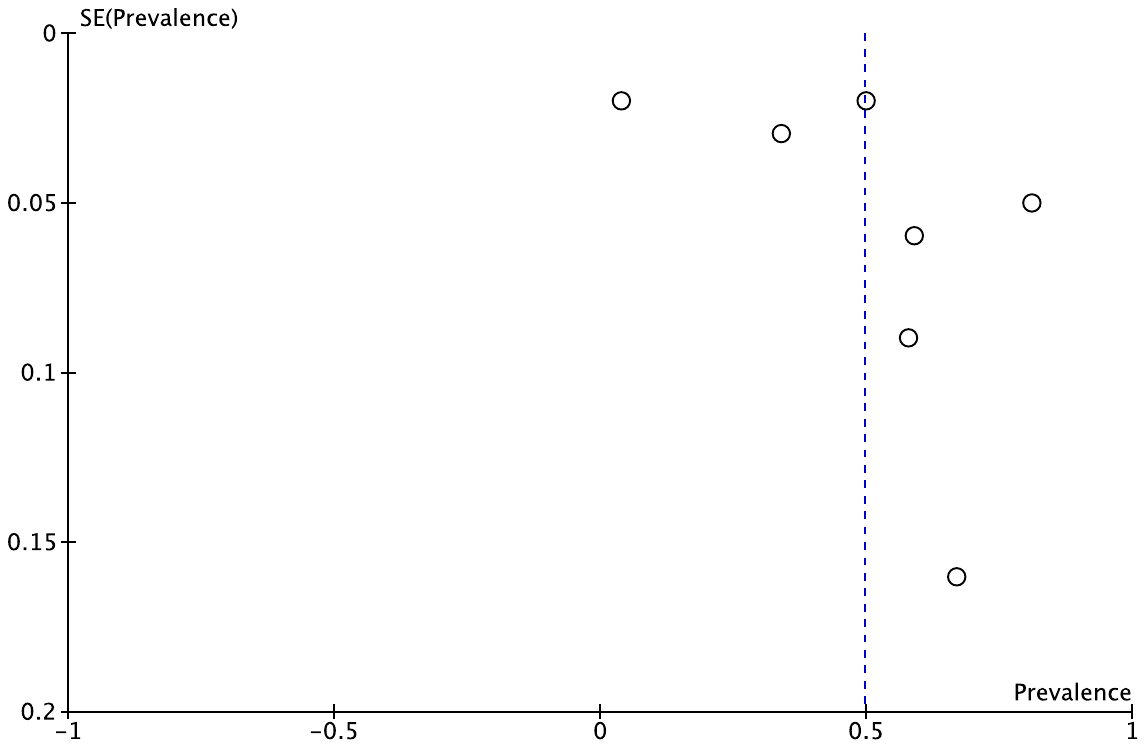

D.

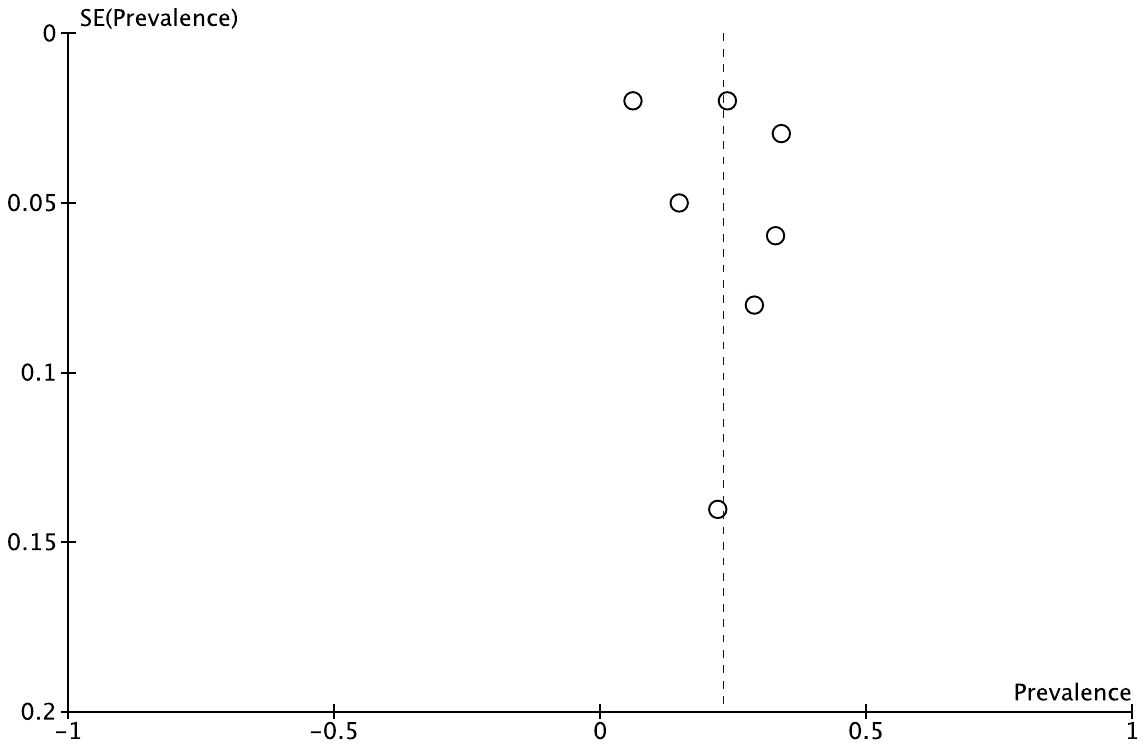

E.

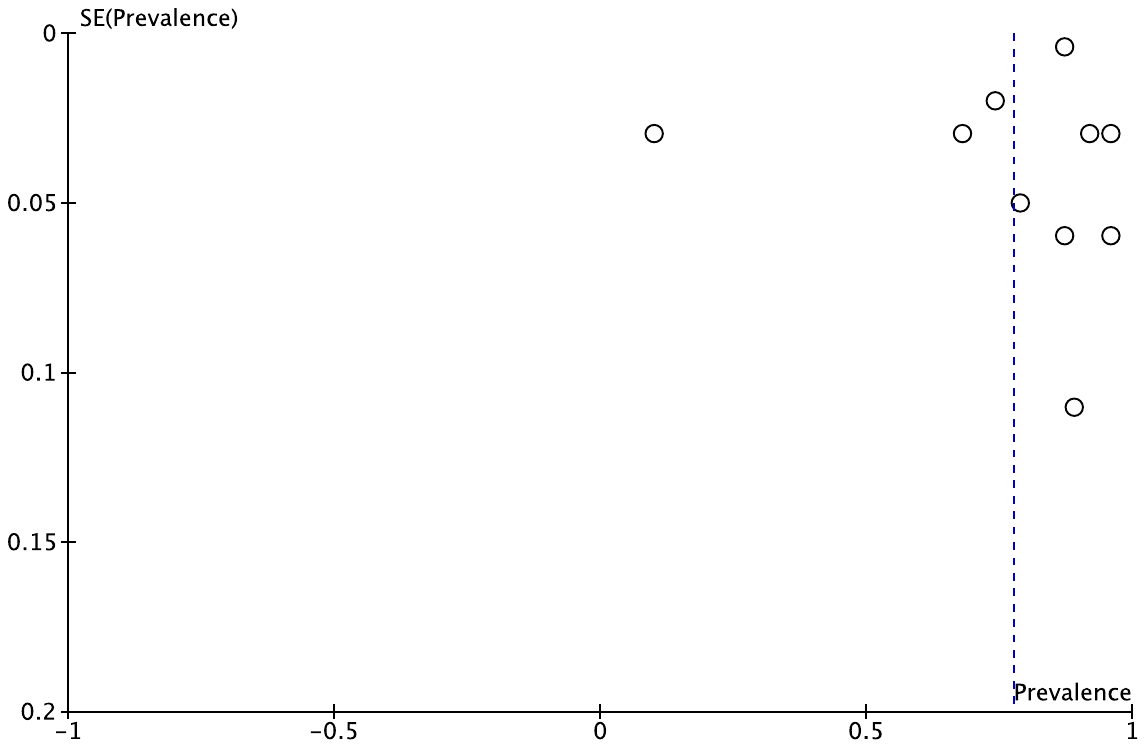

F.

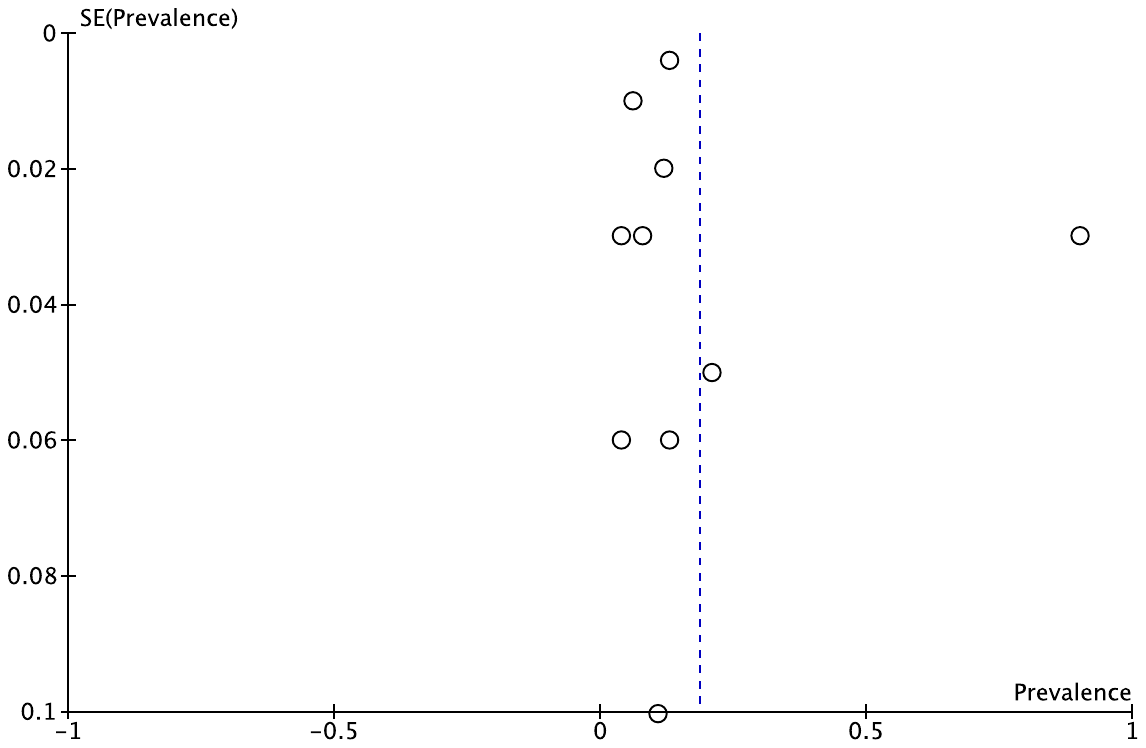

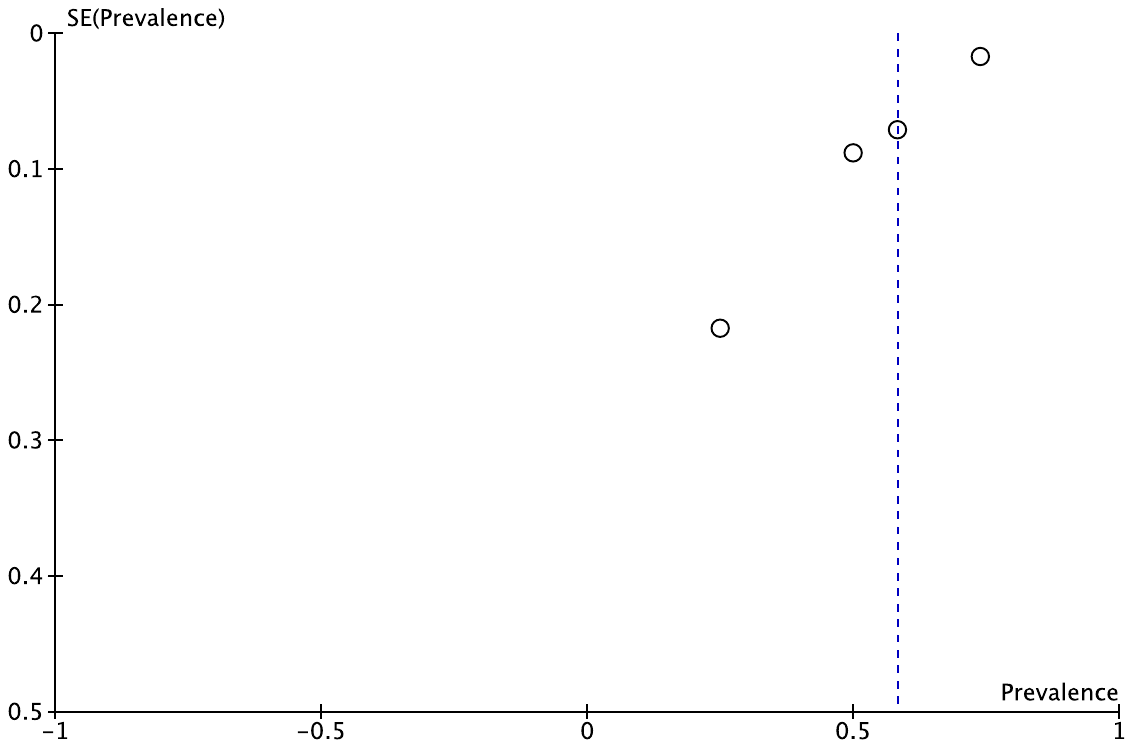
G.

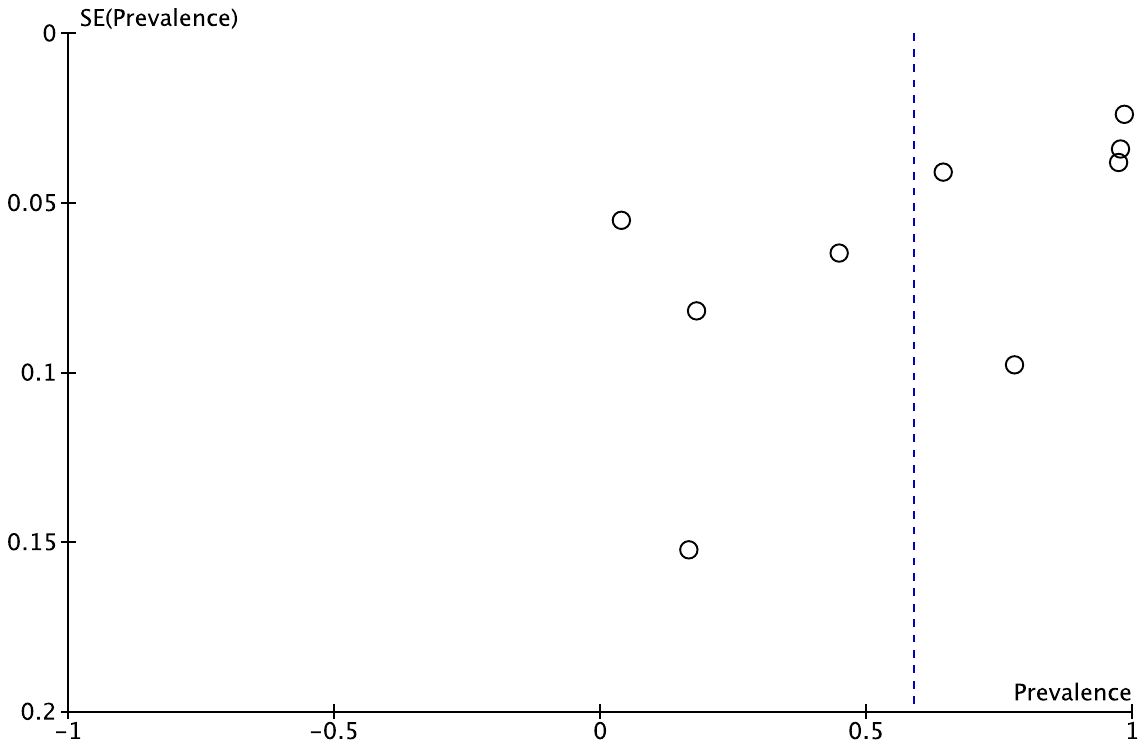
H.

I.
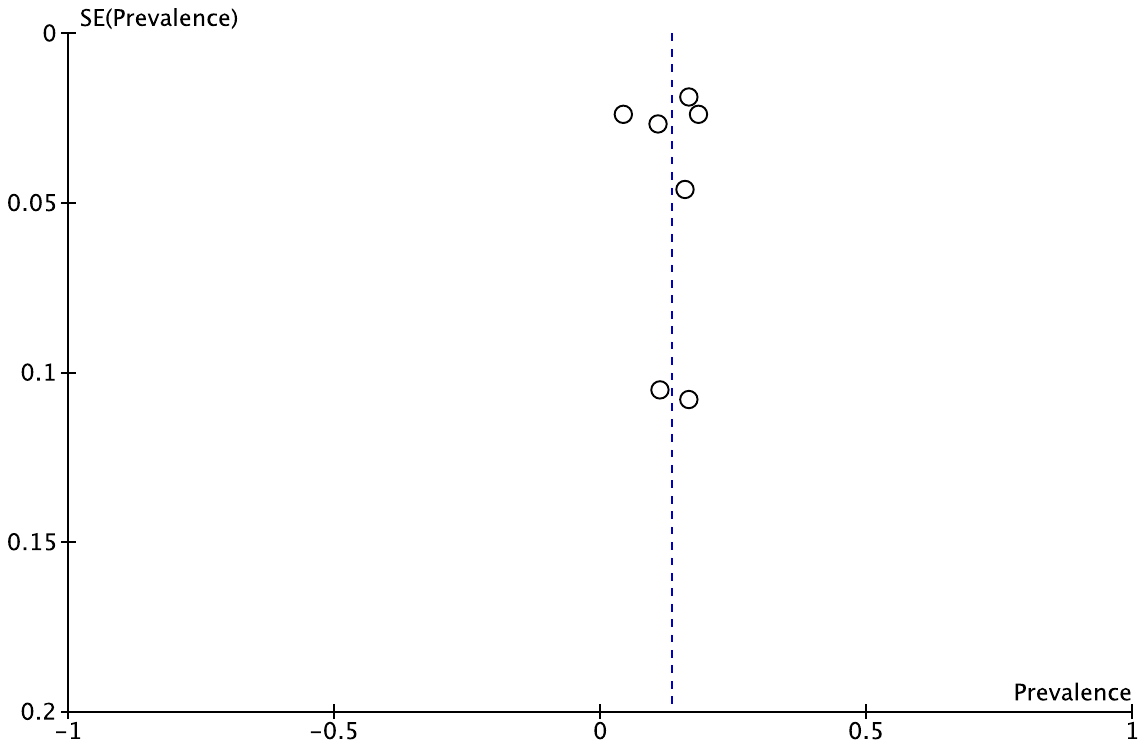

J.

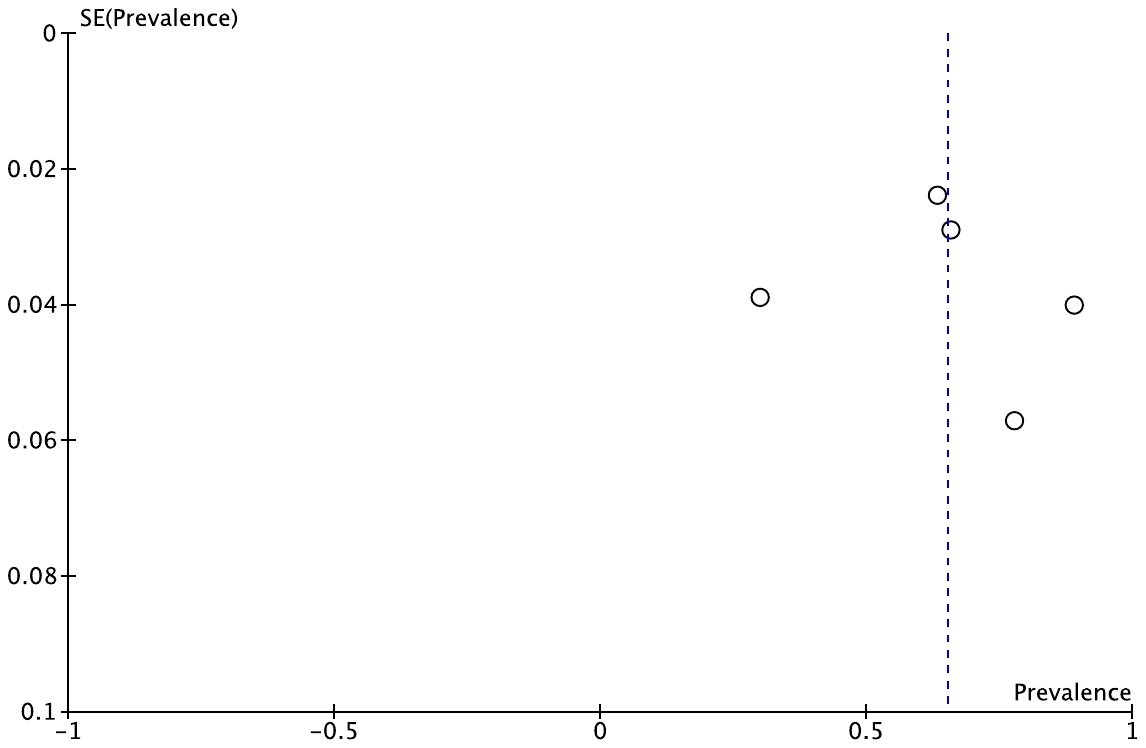

K.

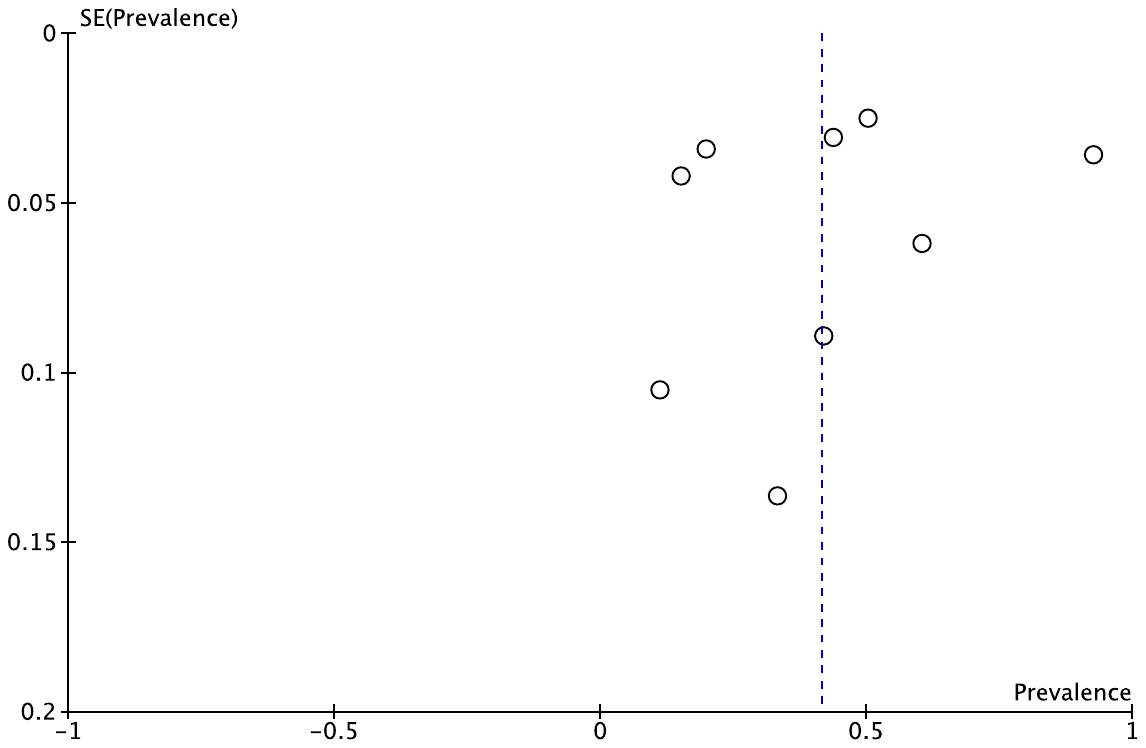

L.

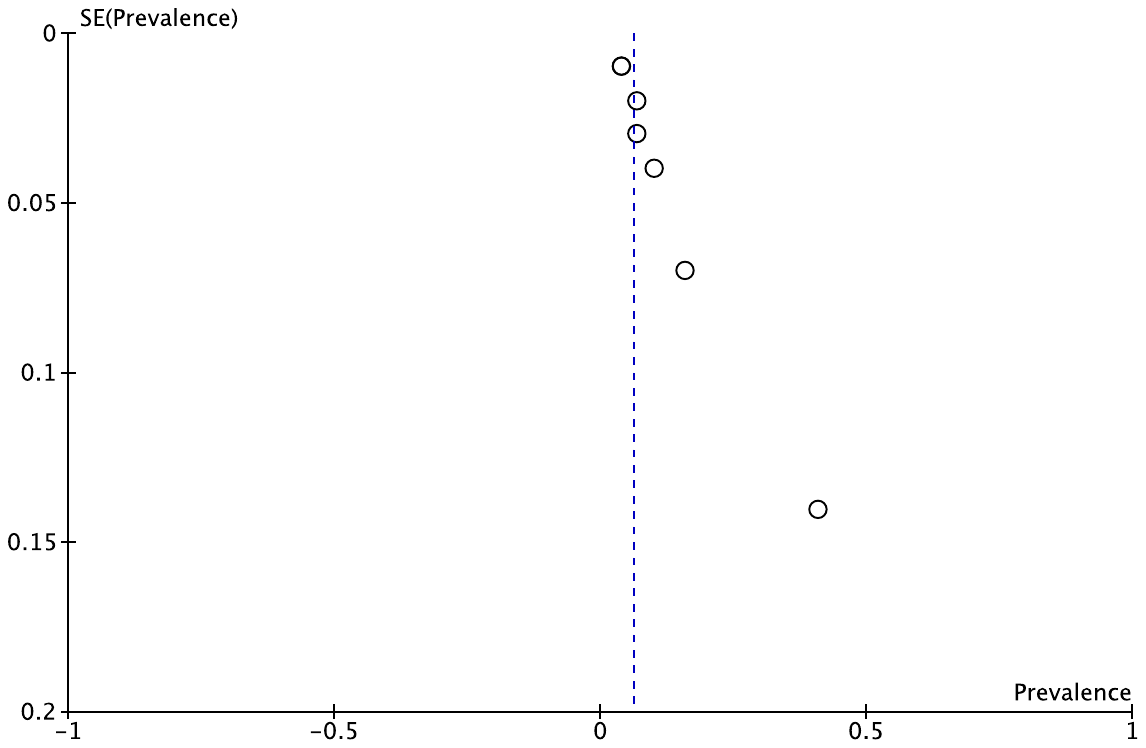

M.

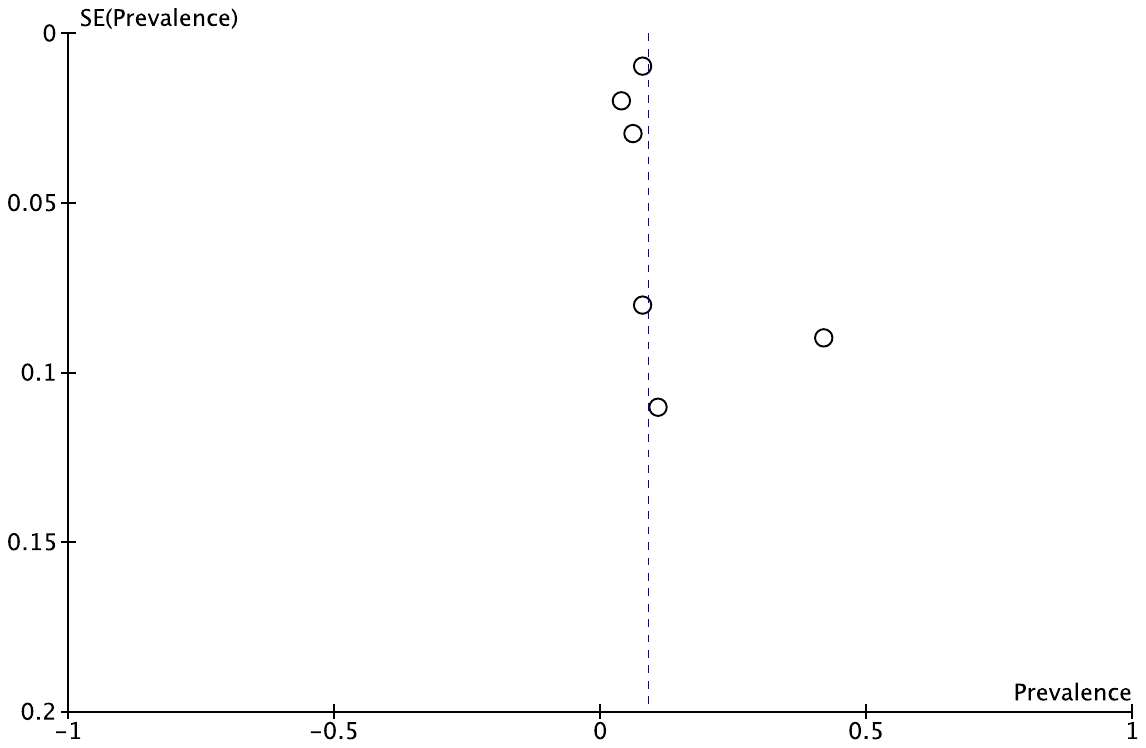

N.
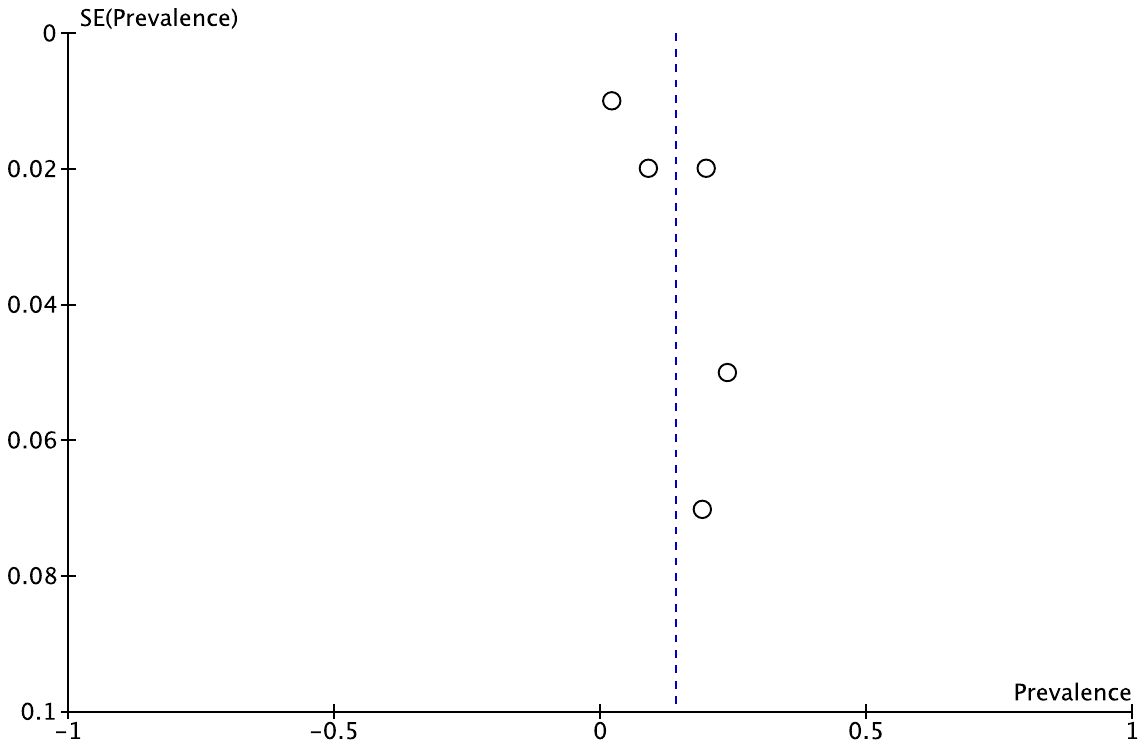

O.

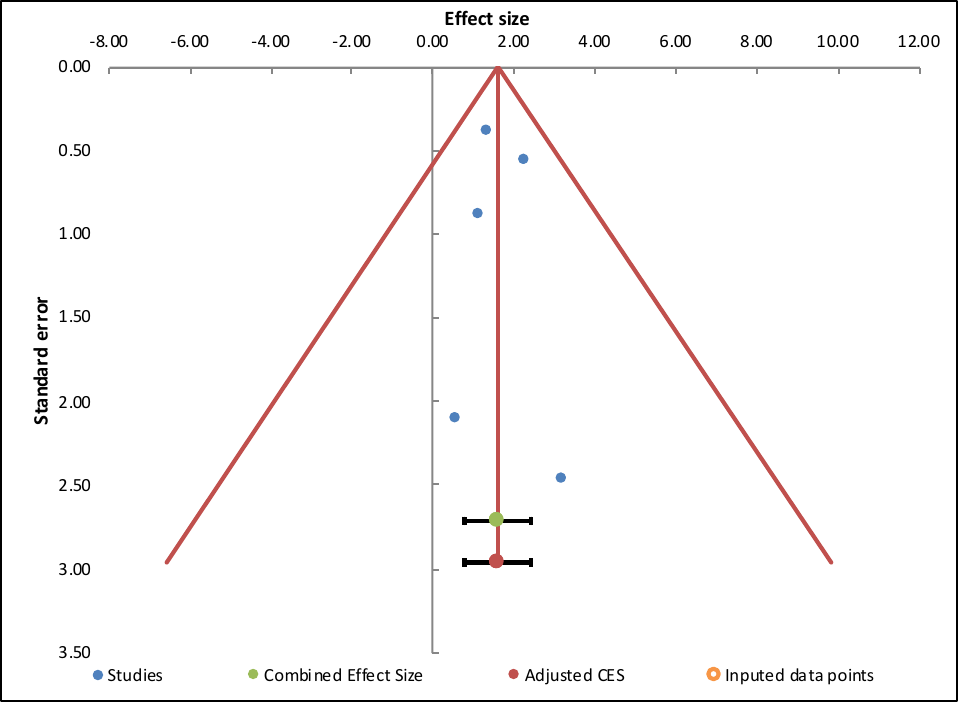

P.

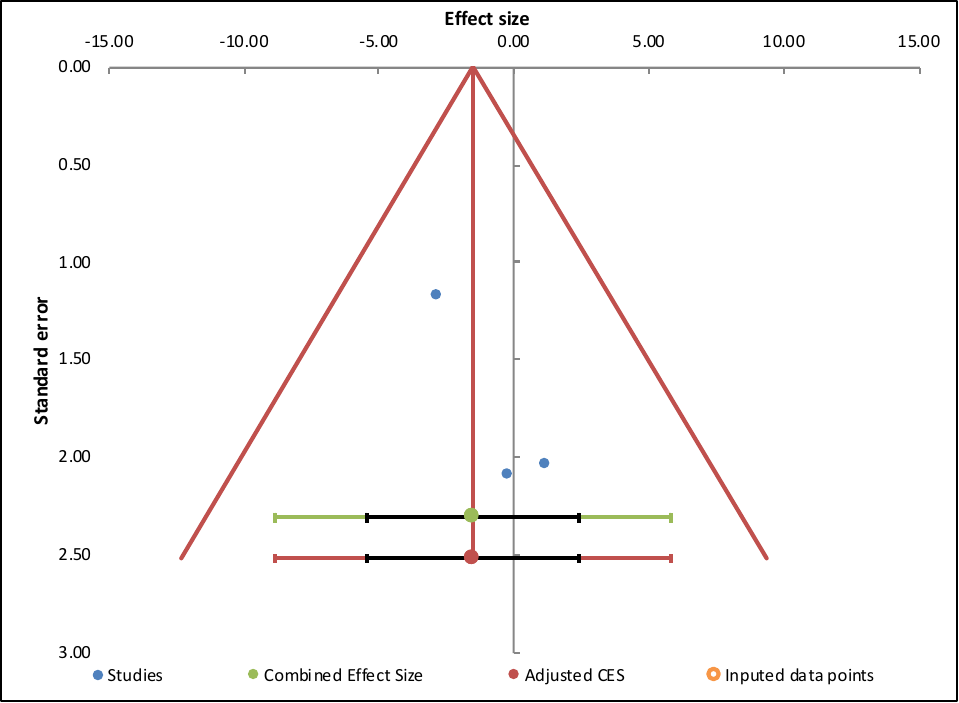
