## Appendix D for "In-hospital cardiac arrest in Intensive Care Unit versus non-Intensive Care Unit patients with COVID-19. A systematic review and meta-analysis"

| Outcome | Egger P value |
| --- | --- |
| Number of IHCAs in hospitalized COVID-19 patients | 0.4 |
| Patients that achieved ROSC | 0.22 |
| Patients with PEA (out of those with IHCA) | 0.37 |
| Patients with asystole (out of those with IHCA) | 0.59 |
| Patients with non-shockable rhythm (out of those with IHCA) | 0.52 |
| Patients with shockable rhythm (out of those with IHCA) | 0.75 |
| Mortality at last follow-up (out of those that achieved ROSC) | 0.17 |
| Patients that had good neurological outcome (CPC 1 or 2) (out of those that survived) | 0.08 |
| Patients with coronary artery disease (out of those with IHCA) | 0.44 |
| Patients with hypertension (out of those with IHCA) | 0.67 |
| Patients with diabetes mellitus (out of those with IHCA) | 0.89 |
| Patients with cancer (out of those with IHCA) | **0** |
| Patients with Chronic Lung Diseases (out of those with IHCA) | 0.2 |
| Patients with Chronic Kidney Diseases (out of those with IHCA) | 0.29 |
| Patients that achieved ROSC (ICU versus general wards) | 0.19 |
| Mortality, out of those with IHCA (ICU versus general wards) | 0.85 |
